# Comparisons of newborn birthweights with maternal factors at Phalombe District Hospital, Malawi: a retrospective record review

**DOI:** 10.1101/2023.09.05.23295074

**Authors:** Dumisani Mfipa, Precious L. Hajison, Felistas Mpachika-Mfipa

## Abstract

**Background:** Birthweight is an important indicator of the newborn’s future health. Maternal factors, including age, HIV status, parity and obstetric complications ([pre]-eclampsia, antepartum hemorrhage [APH] and sepsis), however, have been shown as risk factors of low birthweight (LBW). For data-guided interventions, we compared newborn birthweights with these factors at Phalombe District Hospital, Malawi.

**Methods:** Using a retrospective record review study design, we extracted data of 1,308 women and their newborns from maternity registers (October, 2022-March, 2023). Data were skewed. Its distribution in each group had different variabilities/shapes. We used Mann-Whitney U/Kruskal-Wallis H tests to compare mean rank of birthweights.

**Results:** Prevalence of LBW was 17.4% and median birthweight was 2,900.00g (interquartile range [IQR] 2,600.00g-3,200.00g). We observed significant difference in newborn birthweights among adolescent girls (≤19 years), young women (20-24 years), older women (25-34 years) and women of advanced maternal age (≥35 years), (mean ranks: 600.32, 650.85, 690.62 and 735.34, respectively, H[3] = 20.30, p<.001, η^2^ = 0.01). Pairwise comparisons showed significant differences in newborn birthweights of adolescent girls and older women (p = .006), adolescent girls and women of advanced maternal age (p<.001). We observed no significant differences in newborn birthweights between HIV+ and HIV- women (mean ranks: 608.86 and 659.28, respectively, U = 67,748.50, Z = -1.417, p = .157, r = 0.04). We found significant differences in newborn birthweights between primiparous and multiparous women (mean ranks: 600.95 and 697.16, respectively, U = 180,062.00, Z = -4.584, p<.001, r = 0.13), women with and women with no (pre)-eclampsia, APH and sepsis (mean ranks: 340.09 and 662.64, respectively, U = 10,662.00, Z = -4.852, p<.001, r=0.13).

**Conclusion:** Significant differences reported notwithstanding, small effect sizes and a high prevalence of LBW were observed. Thus, all pregnant women should be prioritized to improve birthweight outcomes. Those with complications, however, require special care.

## Introduction

Birthweight is an important indicator of the newborn’s future health [1]. Prevalence of low birthweight [LBW], however, remains high in resource-constrained countries. World Health Organization (WHO) defines LBW as weight at birth of less than 2,500g, regardless of gestational age and sex [2]. It occurs due to either delivery of preterm or a growth restricted fetus [3]. Of particular importance, LBW is a vital indicator of maternal and child health service performance in a country [2]. Particularly, LBW is the main cause of morbidity and mortality among children aged below 5 years across the world [4,5]. In sub-Saharan Africa (SSA), LBW is a strong determinant of mortality among newborn infants [6]. There are numerous long-term outcomes associated with both preterm and fetal growth restriction, mainly neurologic outcomes that include cerebral palsy, blindness, deafness and hydrocephaly [3].

World Health Assembly in 2012, as a decision-making body of WHO, resolved that LBW prevalence be reduced by a target of at least 30% between 2012 and 2025 as part of the Comprehensive Implementation Plan on Maternal, Infant and Young Child Nutrition [7]. Evidence from studies done elsewhere suggest that birthweights of newborns vary according to maternal age, HIV status, obstetric complications [antepartum hemorrhage, (pre) - eclampsia and sepsis] and parity [8–11]. This is similar to findings from a Malawian study which suggested that birthweights of newborns differ according to maternal HIV status [12].

Unpublished data for Phalombe District Hospital shows that the facility had registered 12% LBW deliveries from 10^th^ October to 31^st^ December 2022 and 9% LBW deliveries from 01^st^ January to 31^st^ March, 2023. This facility was officially opened in October, 2022. We, therefore, compared birthweights of newborns with maternal factors [age, HIV status, parity and obstetric complications {(pre) - eclampsia, antepartum hemorrhage and sepsis}] to understand and to generate local empirical data about newborn birthweight outcomes. The locally-generated empirical evidence will play a key role in designing relevant and targeted interventions to attain the targets set by World Health Assembly [7] and to ensure delivery of quality maternal and child health services at this new facility.

We extracted maternal and newborn data at Phalombe District Hospital covering a period from 10^th^ October, 2022 to 31^st^ March, 2023. We sought to explore the research question ’Do newborn birthweights differ significantly among different categories of maternal age, HIV status, parity and obstetric complications [(pre) - eclampsia, antepartum hemorrhage and sepsis] at Phalombe District Hospital, Malawi?’

## Materials and methods

### Study design and setting

We used a retrospective record review study design. The study was conducted at Phalombe District Hospital in Malawi (Fig. 1).

**Figure 1:**
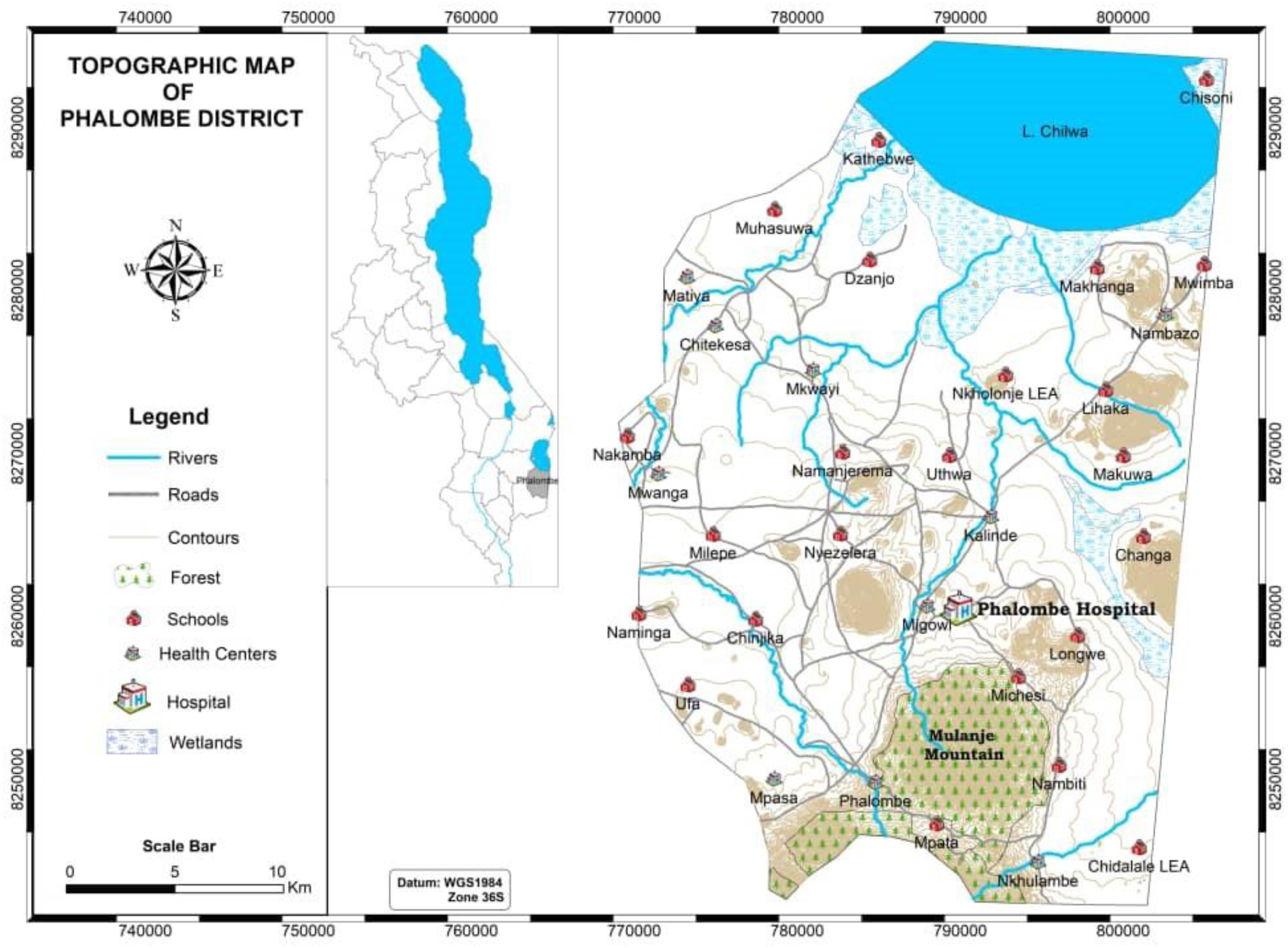
Map of Phalombe District, Malawi.

### Study participants

The study participants were newborns and their respective mothers. The authors had access to maternity registers which had information that could identify individual participants during data collection. However, all data were fully anonymized after data collection such that no single person shall be able to identify individual participants.

### Sample and procedures

All live births from 10^th^ October, 2022 (the first day the facility started offering maternity services) to 31^st^ March, 2023 at Phalombe District Hospital, including those referred from other health facilities were included in the study. Data were accessed between 01^st^ and 10^th^ August, 2023 for research purposes. Data related to newborn birthweights, maternal age, HIV status, parity and obstetric complications [(pre) - eclampsia, antepartum hemorrhage and sepsis] were extracted from the maternity registers.

### Inclusion and exclusion criteria

All live births resulting from singleton pregnancies were included in the study except those newborns with missing birthweights. All fresh stillbirths, macerated stillbirths and neonatal deaths were excluded. In addition, mothers of the sampled newborns with incomplete data were excluded from the study.

### Ethics approval and consent to participate

The research was performed in accordance with the Declaration of Helsinki and written ethical approvals were obtained from the Phalombe District Health Office (DHO) Research Committee and the Malawi National Health Sciences Research Committee (NHRSC) [Approval Number 4131]. This was a retrospective study and both Committees waived the requirement for informed consent.

### Data collection

We developed a data extraction checklist that reflected the components of maternity registers and used it as a guide to extract data from the registers.

### Key variables, definition of variables and measurements

#### a. Independent variables

##### Maternal age

Maternal age was extracted as a continuous variable then categorized and coded as below;19 years and below [adolescent girls] (1), 20-24 years [young women] (2), 25-34 years [older women] (3), 35 years and above [women of advanced maternal age] (4).

##### HIV status

HIV status was categorized and coded as below: HIV negative [refers to women without HIV as confirmed by a new or previous HIV test] (1), HIV positive [refers to women with HIV as confirmed by a new or previous HIV test] (2).

##### Parity

Parity was categorized and coded as below: Primiparous women [women giving birth for the first time] (1) and multiparous women [women giving birth for second time or more] (2).

##### Obstetric complications

This was categorized and coded as below: Absent [refers to women with no (pre) - eclampsia, antepartum hemorrhage and sepsis] (1) and present [refers to women with either (pre) - eclampsia, antepartum hemorrhage and sepsis] (2).

#### b. Outcome variable

Outcome variable was birthweight of newborns measured as a continuous variable in grams.

##### Statistical analysis

Data were entered and analyzed using IBM SPSS Statistics for Windows Version 20.0 (IBM Corp., Armonk, NY, USA). Normality tests by Shapiro-Wilk and Kolmogorov-Smirnov tests were conducted. We concluded that data did not follow a normal distribution. We used box and whisker plots as well as histograms to compare variabilities and shapes of the birthweights’ data among groups, respectively. We observed that the distribution of newborn birthweights in each group had different variabilities and shapes. We, therefore, used Mann-Whitney U test or Kruskal-Wallis H test to compare mean ranks of birthweights. For multiple pairwise comparisons, we reported p-values that were adjusted using the Bonferroni correction. The p-value of less than .05 was considered as the statistical significance for all analyses. In regard to Kruskal-Wallis H test, we calculated the effect size (η^2^) using the eta-squared based on H statistic: eta^2^ [H] = (H - *k* + 1) / (*n*- *k*); where H is the value obtained in the Kruskal-Wallis H test; *k* is the number of groups; *n* is the total of observations [13]. In regard to Mann-Whitney U tests, we calculated effect sizes (*r*) using the formula: Z/√*n;* where Z is the Z statistic; *n* is the number of observations [13]. We interpreted the calculated effect sizes according to criteria by [14]: 0.1 (small effect), 0.3 (moderate effect) and 0.5 (large effect).

## Results

### Maternal factors and newborn birthweight outcomes at Phalombe District Hospital, Malawi

Table 1 shows maternal factors and newborn birthweight outcomes at Phalombe District Hospital in Malawi between October, 2022 and March, 2023. We had 1,308 women (of which 34.4% were adolescent girls) who met the inclusion criteria for this study. Of women with obstetric complications, 63.7% (21/33) had (pre) - eclampsia, 33.3% (11/33) had antepartum hemorrhage and 3.0% (1/33) had sepsis.

**Table 1:** Maternal factors and newborn birthweight outcomes at Phalombe District Hospital in Malawi.

| **Maternal factors** | **N =1,308** | **%** |
| --- | --- | --- |
| Maternal age (in years) |  |  |
| ≤19 | 450 | 34.4 |
| 20-24 | 363 | 27.7 |
| 25-34 | 320 | 24.5 |
| ≥35 | 175 | 13.4 |
| HIV status |  |  |
| HIV- | 1,184 | 90.5 |
| HIV+ | 124 | 9.5 |
| Parity |  |  |
| Primiparous | 580 | 44.3 |
| Multiparous | 728 | 55.7 |
| Obstetric complications† |  |  |
| Absent | 1275 | 97.5 |
| Present | 33 | 2.5 |
| **Newborn birthweight outcomes** |  |  |
| Non - LBW (2,500g and above) | 1,080 | 82.6 |
| LBW (Less than 2,500g) | 228 | 17.4 |

† Obstetric complications in this study refer to (pre) - eclampsia, antepartum hemorrhage and sepsis

### Median birthweight of newborns at Phalombe District Hospital in Malawi

Table 2 shows the mean birthweight of newborns at Phalombe District Hospital in Malawi. Our results show that the median newborn birthweight was 2,900.00g (interquartile range [IQR] 2,600.00g - 3,200.00g) between October, 2022 and March, 2023.

**Table 2:** Median birthweight of newborns at Phalombe District Hospital in Malawi.

| **Newborn birthweight outcomes** | **Total (n = 1,308)** | **Median [IQR]** |
| --- | --- | --- |
| Median birthweight (in grams) | 1,308 | 2,900.00 (2,600.00 - 3,200.00) |

### Normality tests using Kolmogorov-Smirnov and Shapiro-Wilk tests

Table 3 shows the results of the Kolmogorov-Smirnov and Shapiro-Wilk tests to ascertain the distribution of the data. Normality tests conducted by Kolmogorov-Smirnov and Shapiro-Wilk were significant for all categories excluding ’35 years and above’ category of ’maternal age’ variable [Kolmogorov-Smirnov tests (p = .046) and Shapiro-Wilk (p = .061)] and ’present’ category of ’obstetric complications’ variable [Kolmogorov-Smirnov tests (p = .200) and Shapiro-Wilk (p = .940)]. We, therefore, concluded that data did not follow a normal distribution.

**Table 3:** Normality tests using Kolmogorov-Smirnov and Shapiro-Wilk tests.

| Maternal factors | Normality test/Kolmogorov-Smirnov test | P-value | Normality test/Shapiro-Wilk test | P-value |
| --- | --- | --- | --- | --- |
| Maternal age (years) |  |  |  |  |
| ≤19 | 0.071 | **<.001** | 0.982 | **<.001** |
| 20-24 | 0.096 | **<.001** | 0.971 | **<.001** |
| 25-34 | 0.067 | **.001** | 0.985 | **.003** |
| ≥35 | 0.068 | **.046** | 0.985 | .061 |
| HIV status |  |  |  |  |
| HIV- | 0.070 | **<.001** | 0.984 | **<.001** |
| HIV+ | 0.093 | **.010** | 0.977 | **.030** |
| Parity |  |  |  |  |
| Primiparous | 0.073 | **<.001** | 0.983 | **<.001** |
| Multiparous | 0.075 | **<.001** | 0.981 | **<.001** |
| Obstetric complications† |  |  |  |  |
| Absent | 0.070 | **<.001** | 0.984 | **<.001** |
| Present | 0.082 | .200 | 0.986 | .940 |

Bold p-values denote statistical significance at the $p < .05$ level.
† Obstetric complications in this study refer to (pre) - eclampsia, antepartum hemorrhage and sepsis

### Comparisons of newborn birthweights with maternal factors at Phalombe District Hospital in Malawi Maternal age

A Kruskal-Wallis H test showed that there was a statistically significant difference in birthweight of newborns between different maternal age groups, with mean rank birthweights of 600.32 for newborns of adolescent girls (19 years and below), 650.85 for newborns of young women (20-24 years), 690.62 for newborns of older women (25-34 years) and 735.34 for newborns of women of advanced maternal age (35 years and above) [H(3) = 20.30, p < .001, η^2^ = 0.01]. Pairwise comparisons showed a statistically significant difference in birthweight between newborns of adolescent girls and older women (p = .006, adjusted using the Bonferroni correction) as well as adolescent girls and women of advanced maternal age (p < .001, adjusted using the Bonferroni correction). We observed no statistically significant differences in birthweight of newborns between adolescent girls and young women (p = .345, adjusted using the Bonferroni correction) as well as young women and older women (p = 1.000, adjusted using the Bonferroni correction). Data showed no statistically significant differences in birthweight of newborns between young women and women of advanced maternal age (p = .089, adjusted using the Bonferroni correction) as well as older women and women of advanced maternal age (p = 1.000, adjusted using the Bonferroni correction).

### HIV status

A Mann-Whitney U test showed no statistically significant difference in birthweights of newborns between different categories of maternal HIV status, with a mean rank birthweight of 608.86 for newborns of HIV positive mothers and 659.28 for those of HIV negative mothers (U = 67,748.50, Z = -1.417, p = .157, r = 0.04).

### Parity

A Mann-Whitney U showed that there was a statistically significant difference in birthweight of newborns between different categories of maternal parity, with mean rank birthweights of 600.95 for newborns of primiparous mothers and 697.16 for newborns of multiparous mothers (U = 180,062, Z = -4.584, p < .001, r = 0.13).

### Obstetric complications

A Mann-Whitney U test showed that there was a statistically significant difference in newborn birthweights between women with and women with no (pre) - eclampsia, antepartum hemorrhage and sepsis (mean ranks: 340.09 and 662.64, respectively, U = 10,662.00, Z = -4.852, p<.001, r = 0.13).

## Discussion

This study was aimed at comparing birthweights of newborns with maternal factors [age, HIV status, parity and obstetric complications {(pre) - eclampsia, antepartum hemorrhage and sepsis}].

### Maternal age

Our study reported a statistically significant difference in newborn birthweights among different maternal age groups. Specifically, we observed significant differences in newborn birthweights of adolescent girls and older women as well as those of adolescent girls and women of advanced maternal age. We further observed that as the mean ranks of newborn birthweights increased, so was the maternal age group. This is consistent with findings from an Ethiopian study which suggested that maternal age was a protective factor for LBW as an increase in age by one year resulted in a 4% risk reduction (AOR = 0.96, 95% CI 0.92, 1.00) [15]. Similarly, an observational sub Saharan Africa multi-country study suggested that young maternal age increases the risk for adverse pregnancy outcomes and it is also a strong determinant of LBW among newborn infants [6]. In addition, a Bangladesh study reported that young pregnant women (19 years and below) and adult pregnant women (35 years and above) were at increased risk of different adverse birth and health outcomes [16]. Likewise, maternal age at delivery of less than 18 years and above 35 years old was associated with increased risk of LBW [17]. An Ethiopian study on the effects of maternal age and parity on birthweight of newborns of mothers with term and singletons, however, observed that women with a maternal age of 40 and above were associated with a higher risk of delivering LBW newborns compared to a maternal age of 30-34 [8]. The study reported the following mean birthweights of newborns; 3,159.1g (SD = 518.8), p = .765 (19 years and below) vs 3,144.1g (SD = 514.8), p = .718 (20-24 years) vs 3,168.1g (SD = 506.2), p = .299 (25-29 years) vs 3,004.2g (SD = 589.9), p < .001 (30-34 years) vs 2,844.7g (SD = 664.3), p = .052 (35-39 years) vs 2,397.8g (SD = 673.5), p = .006 (40 years and above) [8]. Slightly over one-thirds (34.4%) of the women in our study were adolescent girls. This is consistent with previous findings which showed that 29% of adolescent girls (15-19 years) have begun child bearing in Malawi [18]. We, therefore, encourage both state and non-state actors to advocate against child marriages and adolescent pregnancies. The higher number of LBW babies in adolescents in this study can further be attributed to biological immaturity as the adolescent is still growing and developing and may compete for nutrient with the unborn baby [19]. Hence, we further recommend that adolescent SRHR services should be scaled up to increase access to and usage of contraceptives to deter adolescent pregnancies and child bearing among adolescent girls which contribute significantly to poor birthweights of newborns at Phalombe District Hospital. Importantly, efforts to improve birthweight outcomes should target all pregnant women regardless of their maternal age group as the effect size reported in our study was small.

### HIV status

We observed that there were no statistically significant differences in mean ranks of newborn birthweights born to HIV positive and HIV negative women. This is in contrast with findings from a previous Malawian study that analyzed DHS data for 2010 which found that the mean birthweight of infants born to HIV positive mothers was statistically significantly lower compared to infants born to HIV negative mothers (3,192g [SD = 724] vs 3,275g [SD = 723], p =. 0412) [12]. Likewise, the frequency of LBW infants among HIV negative mothers was statistically significantly lower from their HIV positive counterparts (9.26% vs 13.6%, p = .0337) [12]. This finding in our study can be attributed to a robust, well-coordinated and effective Prevention of Mother to Child Transmission (PMTCT) of HIV Option B+ program, which has greatly improved health outcomes of newborns and mothers over the years in Malawi in general and in Phalombe district in particular. We, therefore, encourage health authorities to intensify PMTCT programs in the district, including in hard-to-reach areas to sustain the achievements made and maximize the benefits this program provides to HIV exposed newborns and HIV infected mothers. We, also, urge health authorities to continue sensitizing pregnant women on the importance of early antenatal care (ANC) as this offers an opportunity to ascertain the HIV status and linkage to care in good time hence the improved health status of the mother and prevention of possible adverse effects to the newborn, including low birthweight [20].

### Parity

Our results showed that newborns born to primiparous mothers had a significantly lower mean rank birthweight as compared to those born to multiparous mothers. This finding is similar to a Polish study found that the mean birthweights of neonates born to primiparous mothers and multiparous mothers were 3,356.3g (SD = 524.9) and 3,422.7g (SD = 538.6), respectively (p ≤ .001) [11]. Similarly, a different Polish study and an India study observed that primiparous mothers had a statistically significant higher frequency of LBW than multiparous mothers [21,22]. This is in contrast to the findings from an Ethiopian study which observed that primiparous women had less risk of having an LBW baby compared to multiparous women [8]. The difference in newborn birthweights between primiparous and multiparous mothers in the present study could be due to the physiological changes that occur in subsequent pregnancies as compared to primigravidas hence the known fact that primiparity increases the risk of LBW [23]. Biologically, among other factors, the uteroplacental blood flow is lesser in first pregnancy compared to subsequent pregnancies and structural factors which limit uterine capacity are present in first pregnancy [24]. We suggest that during antenatal care, women with first pregnancy should be offered all the interventions in the antenatal care package to prevent increasing the likelihood of a LBW baby.

### Obstetric complications

We found that newborns of women with (pre) - eclampsia, antepartum hemorrhage and sepsis had a significantly lower mean rank birthweight as compared to those of women with no (pre) - eclampsia, antepartum hemorrhage and sepsis. This is similar to a prospective cohort study in a tertiary referral center in urban Uganda that observed that mean birthweight for pre - eclampsia cases was 2.48 kg (SD = 0.81) compared to 3.06 kg (SD = 0.46) for controls (p < .001) [9]. We encourage health facilities to prioritize as well as intensify comprehensive physical examination and screening for obstetric complications, including (pre) - eclampsia, antepartum hemorrhage and sepsis among pregnant women attending ANC services to prevent development of these complications and identify the high-risk cases and offer appropriate management. In this regard, lessons can be drawn from a study on utilization of cervical cancer screening (CCS) services which observed that the proportion of women who had done CCS was significantly higher among those who were recommended by health workers than those who were not [25]. Patient-centered care and exclusion of pregnancy related complications like (pre) - eclampsia, antepartum hemorrhage and sepsis may afford many pregnant women an opportunity to receive quality care before delivery and improve birthweights of newborns in the district. Of note is the fact that only 3.0% (1/33) of the reported obstetric complications were sepsis cases in our study. This is a good observation and feedback to the Phalombe District Hospital staff as both intra and post-partum sepsis relates to the quality of care during labour and delivery. Despite the significant differences in newborn birthweight between women with and women with no (pre) - eclampsia, antepartum hemorrhage and sepsis, we recommend that every pregnant woman should be provided with quality maternal care services to improve newborn birthweights in view of the small effect sizes reported.

### Limitations of the study

This was a retrospective record review study design as such we relied on data that was recorded in maternity registers with some variables having incomplete and missing data. This led to the exclusion of some mothers and their newborns from the study. Nonetheless, the large sample of the data that was collected and analyzed compensated for those that did not meet the inclusion criteria. Our selection of independent variables was also guided by and restricted to information that was available in the maternity registers. This meant that we could not analyze socio- demographic data of the mothers in our study as these registers do not capture such data. The results of this study should be generalized with caution as the study was conducted at one facility in the district. However, this is a referral hospital for the district such that it was highly likely that newborns of mothers referred from other facilities within the district were represented in the study. Our study included data for each and every preterm, term and postterm newborn as long as they met the inclusion criteria. Hence, it is important to generalize these results with caution.

## Conclusions

In this study, the research question ’Do newborn birthweights differ significantly among different categories of maternal age, HIV status, parity and obstetric complications at Phalombe District Hospital, Malawi?’ was explored. Our results revealed statistically significant differences in newborn birthweights between adolescent girls and older women, adolescent girls and women of advanced maternal age, women with and women with no obstetric complications as well as primiparous and multiparous women. Our study showed no statistically significant difference in newborn birthweights between HIV+ and HIV- women. We also observed a high prevalence of LBW. This research has provided important insights about newborn birthweights between different maternal factors at this facility. It is, however, important that future research should focus on comparing newborn birthweights with socio-demographic data and identifying the predictors of LBW at this facility. The future research would contribute to a more comprehensive understanding of newborn birthweights between different socio-demographic factors and risk factors of LBW at this facility. Our study contributes to the field of maternal, newborn and child health as it is builds upon prior studies and enhances the understanding and the role that maternal factors play regarding newborn birthweight outcomes. This study establishes statistically significant differences in newborn birthweights between different maternal age, parity and obstetric complication. Our study, however, supports the evidence that an increase in maternal age could be a protective factor for LBW as it resulted in LBW risk reduction. It is, however, important to note that our study observed small effect sizes. We, therefore, advise implementers at district and community levels to prioritize interventions aimed at improving birthweight outcomes for all groups of pregnant women, including preventing adolescent pregnancies, adolescent childbirths, (pre) - eclampsia, antepartum hemorrhage, sepsis and pay attention to the needs of primiparous women.

## List of abbreviations

ANC: antenatal care
APH: antepartum hemorrhage
CEmONC: comprehensive emergency obstetric and newborn care
HIV: human immunodeficiency virus
IQR: interquartile range
LBW: low birthweight
PMTCT: prevention of mother to child transmission of HIV.

## Declarations

### Consent for publication

Not applicable

### Availability of data and materials

The datasets generated during and/or analyzed during the current study are available from the corresponding author on a reasonable request.

### Authors’ contributions

DM conceived and designed the study, analyzed the data and prepared the draft manuscript. All authors reviewed the manuscript, read and approved the final manuscript.

## Acknowledgments

We are greatly indebted to Hassan Mdala for designing the study map and Phalombe District Health Office (DHO) for granting us permission to conduct the study at their health facility.

## References

1. Wubetu AD, Amare YE, Haile AB, Degu MW. Newborn Birth Weight and Associated Factors Among Mother-Neonate Pairs in Public Hospitals, North Wollo, Ethiopia. Pediatr Health Med Ther. 2021;12:111–8.

2. World Health Organization [WHO]. The WHO application of ICD-10 to deaths during the perinatal period: ICD-PM. WHO. 2016;

3. Goldenberg RL, Culhane JF. Low birth weight in the United States. Am J Clin Nutr. 2007;85(2):584–90.

4. World Health Organization [WHO]. Preterm birth [Internet]. WHO; 2022 [cited 2023 Jan 18]. Available from: https://www.who.int/news-room/fact-sheets/detail/preterm-birth

5. de Bernabe JV, Soriano T, Albaladejo R, Juarranz M, Calle ME, Martinez D, et al. Risk factors for low birth weight: a review. Eur J Obstet Gynecol Reprod Biol. 2004;116(1):3–15.

6. Mombo - Ngoma G, Mackanga JR, Gonzalez R, Ouedraogo S, Kakolwa MA, Basra A, et al. Young adolescent girls are at high risk for adverse pregnancy outcomes in sub - Saharan Africa: an observational multi-country study. BMJ Open. 2016;6(6):e011783.

7. World Health Organization [WHO]. Comprehensive Implementation Plan on Maternal, Infant and Young Child Nutrition. WHO. 2014;

8. Bekele A, Seyoum G, Tesfaye K, Fantahum Y. The effects of maternal age and parity on the birth weight of newborns among mothers with singleton pregnancies and term deliveries. Ethiop J Health Dev. 2019;33(3):182–7.

9. Nakimuli A, Starling JE, Nakubulwa S, Namagembe I, Sekikubo M, Nakabembe E, et al. Relative impact of pre-eclampsia on birth weight in a low resource setting: A prospective cohort study. Pregnancy Hypertens. 2020;21:1–6.

10. Maharlouei N, Mansouri P, Zahmatkeshan M, Lankarani KB. Low-risk planned caesarean versus planned vaginal delivery at term: early and late outcomes. East Mediterr Health J. 2018;25(7):503–13.

11. Genowska A, Motkowski R, Strukcinskaite V, Abramowicz P, Konstantynowicz J. Inequalities in Birth Weight in Relation to Maternal Factors: A population - Based Study of 3, 813, 757 Live Births. Int J Environ Res Public Health. 2022;19(3):1384.

12. Msamila S. “The Association between Maternal HIV Status and Low Birth Weight Offspring, Malawi DHS 2010” Thesis [Internet]. [Georgia]: Georgia State University; 2018. Available from: 10.57709/11017317

13. Tomczak M, Tomczak E. The need to report effect size estimates revisited. An overview of some recommended measures of effect size. Trends Sport Sci. 2014;1(21):19–25.

14. Cohen J. Statistical Power Analysis for the Behavioral Sciences. 2nd ed. Hillsdale, NJ: Lawrence Erlbaum Associates, Publishers; 1988.

15. Vahdaninia M, Tavafian SS, Montazeri A. Correlates of low birth weight in term pregnancies: a retrospective study from Iran. BMC Pregnancy Childbirth. 2008;8(12):1–5.

16. Alam R, Khan N, Cherri Z, Tapan Kumar R, Rahman M. Child Bearing Age and Pregnancy Outcomes in Bangladesh: A Multilevel Analysis of a Nationwide Population-Based Survey. Prim Health Care. 2018;8(2):2–5.

17. Momeni M, Danaei M, Kermani A, Bakhshandeh M, Foroodnia S, Mahmoudabadi Z, et al. Prevalence and Risk Factors of Low Birth Weight in the Southeast of Iran. Int J Prev Med. 2017;8:12.

18. National Statistical Office [NSO] Malawi and ECF. Malawi Demographic and Health Survey 2015-16. Zomba, Malawi, and Rockville, Maryland, USA: NSO and ICF; 2017.

19. Restrepo-Méndez MC, Lawlor DA, Horta BL, Matijasevich A, Santos IS, Menezes AM, et al. The Association of Maternal Age with Birthweight and Gestational Age: A Cross-Cohort Comparison. Paediatr Perinat Epidemiol. 2015;29(1):31–40.

20. Kapito-Tembo AP, Bauleni A, Wesevich A, Ongubo D, Hosseinipour MC, Dube Q, et al. Growth and Neurodevelopment Outcomes in HIV-, Tenofovir-, and Efavirenz-Exposed Breastfed Infants in the PMTCT Option B+ Program in Malawi. J Acquir Immune Defic Syndr. 2021;86(1):81–90.

21. Merklinger - Gruchala A, Jasienska G, Kapiszewska M. Paternal investment and low birth weight – The mediating role of parity. PLOS ONE. 2019;14(1):e0210715.

22. Kaur J, Kaur K. Obstetric complications: Primarity vs. Multiparity. Eur J Exp Biol. 2012;2(5):1462–8.

23. Best R, Mvala J, Dube A, Katundu C, Kalobekamo F, Mortimer K, et al. A secondary data analysis of a cluster randomized controlled trial: improved cookstoves associated with reduction in incidence of low birthweight in rural Malawi. Int J Epidemiol. 2022;51(6):1803– 12.

24. Hinkle S, Albert P, Mendola P, Sjaarda L, Yeung E, Boghossian N, et al. The association between parity and birthweight in a longitudinal consecutive pregnancy cohort. Paediatr Perinat Epidemiol. 2014;28(2):106–15.

25. Mpachika-Mfipa F, Kululanga LI, Mfipa D, Kazembe A. Utilization of cervical cancer screening and its associated factors among women of child-bearing age in Mangochi district, Malawi: a facility-based cross-sectional study. BMC Women’s Health. 2023;334.

